# Hospital profiling using Bayesian decision theory

**DOI:** 10.1101/2021.06.23.21259367

**Authors:** Johannes Hengelbrock, Johannes Rauh, Jona Cederbaum, Maximilian Kähler, Michael Höhle

## Abstract

**Background:** For evaluating the quality of care provided by hospitals, special interest lies in the identification of performance outliers. We study a setting where the decision to classify hospitals as performance outliers or non-outliers is based on the observed result of a single binary quality indicator.

**Methods:** We propose to embed the classification of providers into a Bayesian decision theoretical framework which enables the derivation of optimal decision rules with respect to the expected decision consequences. We argue that these consequences depend upon for which pathway to quality improvement the profiling of hospitals takes place. We propose paradigmatic utility functions for the two pathways *external reporting* and *change in care delivery* and compare the resulting optimal decision rules with regard to their threshold values, sensitivity and specificity. We further apply them to the area of hip replacement surgeries by analyzing data from the mandatory German hospital profiling program. Based on five quality indicators, we re-evaluate the performance of 1,277 hospitals which treated over 180,000 patients for hip-replacement surgeries during 2017.

**Results:** Based on the utilities we assigned to the classification decisions, the decision rule for change in care delivery classifies more high-volume providers as outliers compared to the decision rule for external reporting, with consequences for both sensitivity and specificity. The re-evaluation of the five quality indicators illustrates that classification decisions are highly dependent upon the underlying utilities.

**Conclusion:** Analyzing the classification of hospitals as a decision theoretic problem and considering pathway-specific consequences of decisions can help to derive an appropriate decision rule. Contrasting decision rules with regard to their underlying assumptions about the utilities of classification consequences can be helpful to make implicit assumptions transparent and justifiable.

## Introduction

The evaluation of the quality of care that patients receive from hospitals or other healthcare providers has gained increasing public and political attention (1,2). Measurements of quality of care are used for *external reporting* as well as for initiating *change in care delivery* (in the follwing referred to as *change*) with the common aim of quality improvement (3)^1^. For these purposes, special interest lies in the identification of performance outliers, either compared to a national average or a pre-specified target (5,6). For estimating and classifying hospital performances based on observed results, various frequentist and Bayesian statistical methods have been proposed and compared (5,7–10). However, little attention has been paid to the question of how much statistical evidence is needed in order to classify a hospital as performance outlier. Often, simply a pre-specified level of statistical significance is used as classification threshold, usually without relation to the consequences of the classification decision (9,11).

As an alternative, previous studies suggested to specify utility functions that quantify the potential costs or utilities associated with classification decisions (12–14). These include a generalized version of a 1-0 utility function for classification decisions (13) but also more complex ones that take into account the consequences from different stakeholder perspectives (14). We extend this work by analyzing and comparing the consequences of two different utility functions: a generalized 1-0 utility function for external reporting and a utility function that depends upon the underlying quality of care as well as the number of affected patients for change in care delivery. Using an exemplary quality indicator for a proportion, we use funnel plots to illustrate how the optimal decision rule depends upon the choice of a utility function and analyze the consequences for the sensitivity and specificity of the classification decision in different scenarios and as a function of the number of treated patients. In order to illustrate the practical consequences, we then apply both utility functions to data on hip replacement surgeries that was collected for hospital profiling in Germany during 2017 and re-evaluate five indicators of quality of the mandatory German hospital profiling program. These indicators cover over 180,000 treated patients in 1,277 hospitals. Finally, we discuss the insights such an formal decision theoretic view can provide for the practical process of the evaluation of quality of care, as well as its limitations.

## Methods

As starting point, we assume that a binary *quality indicator* is used to measure a certain aspect of the quality of care provided by a hospital (or some other healthcare provider), for which data is collected and analyzed. Typical examples include the adherence to specified processes (15,16) or outcome indicators such as 30-day mortality after some procedure (17). Data is collected for a pre-specified time period and the population of interest are all patients fulfilling certain inclusion criteria defined for the indicator within the selected time period. In the following, *O*_*j*_ denotes the binary outcome of interest of patient *j, j* = 1, …, *J*, at a specific hospital. In the case of an annual evaluation of the quality of care, for instance, *J* is the number of patients which are considered as part of the quality indicator in the year of interest. We assume that the outcome variables are independent and identically distributed Bernoulli variables with outcome probability P(*O*_*j*_ = 1) = *θ*. This implies that individual patient characteristics are assumed to have no relevant effect on the outcome of interest. Our framework can however be extended to risk adjusted quality indicators with patient-specific event probabilities. In the following, *O* denotes the sum of the outcome variables of interest, 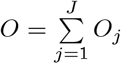, that is the number of events of interest at the specific hospital. By assumption, *O*|*J, θ* ∼ Bin(*J, θ*).

In the case of non-risk adjusted quality indicators, the evaluation of hospital performance is based on the comparison of an estimate of the outcome probability *θ* with one or multiple pre-specified target values, which are the same for all providers. In our work, we focus on the case of binary classification based on a single pre-specified target value *R*, but the framework can easily be extended to multiple categories. In the binary case, hospitals are often classified into *performance outliers* and *non-outliers*, respectively (9). Furthermore, we assume that the target value *R* is the upper bound of a reference range and that the goal is to identify hospitals with true underlying values of *θ* above the target as performance outliers. We assume in the following that the target value *R* is a specified value known before the start of the data collection (2). Alternatively, *R* can also be an estimated mean or some quantile of the observed indicator results of several providers (5). Interest lies in the underlying value *θ* (opposed to just the observed rate *o/J* with *O* = *o*), because we are interested in the inherent quality processes at the hospital. This view corresponds to a so called analytic analysis framework in the spirit of (18).

The classification decision dividing hospitals into performance outliers and non-outliers can be conceptually formulated as an influence diagram (19,20), in which nodes represent random variables, arrows indicate conditional dependencies of the joint probability distribution and thus the absence of arrows indicates conditional independence. Figure 1 illustrates the case of a non-risk adjusted quality indicator operationalized as a proportion:

**Figure 1:**
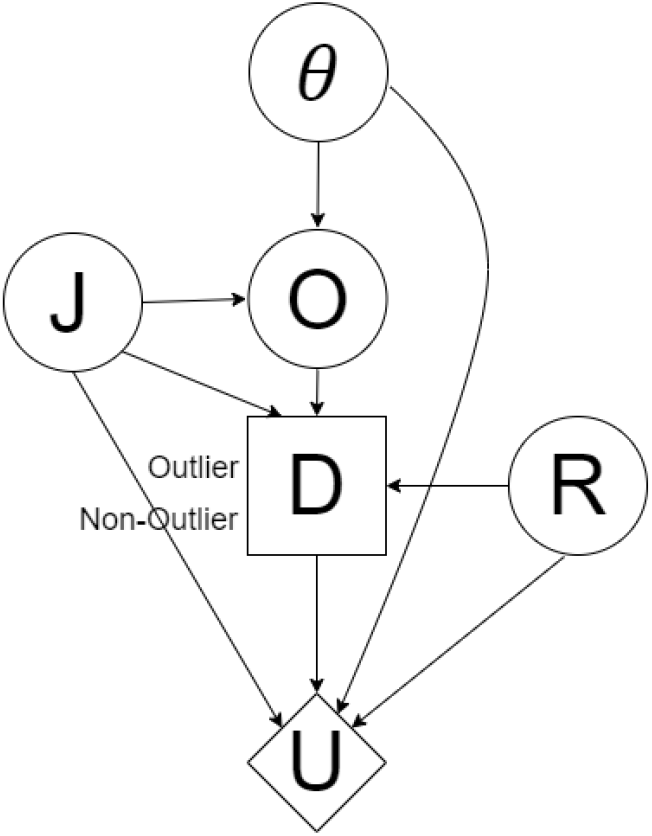
Influence diagram for the classification of a single hospital

*D*(·) denotes the decision to classify a hospital into one of the two categories *D* ∈ {outlier, non-outlier}, and *U* represents the utility function. Note that the number of patients *J* and the latent parameter of interest *θ* are assumed to be conditionally independent; this assumption can be relaxed, if necessary. From Figure 1, it follows that the classification decision depends upon the parameter of interest *θ*, the number of treated patients *J*, the observed number of outcomes of interest, the target value *R* as well as the utility function.

In a Bayesian context, for all random variables without parent nodes in the graph, a prior distribution has to be specified, reflecting all relevant prior knowledge. Since the decision *D* takes into account *R* and *J*, we may assume that *R* and *J* are not random. Thus, formally, we use point mass priors for *R* and *J*. Therefore, only a prior distribution for *θ* is really needed.

Given a treated number of patients *J* and an observed number of events of interest *O* = *o*, an optimal decision rule for the classification of a single hospital can be derived by maximizing the expected utility associated with the two classification decisions. This leads to a Bayes optimal decision rule (21):

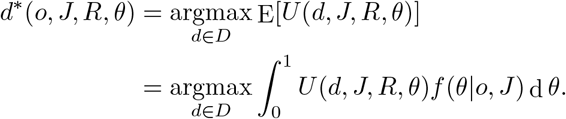

Here, *f* (*θ*|*o, J*) denotes the density of the posterior distribution of *θ* given *o, J* and a prior distribution of *θ*.

For the simultaneous classification of multiple providers *i* = 1, …, *I*, the Bayes optimal decision rule is a vector of length *I*, denoted as ***d***^*∗*^(). For multiple providers, we assume an additive utility function

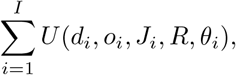

with the expected utility of each individual decision satisfying E[*U* (*d*_*i*_, *o*_*i*_, *J*_*i*_, *R, θ*_*i*_)] ≥ 0. Additivity of utilities is a simplifying assumption that does not necessarily hold in all decision contexts. In the discussion, we refer to situations in which simple additivity may not suitable.

The Bayes optimal decision rule is then defined as

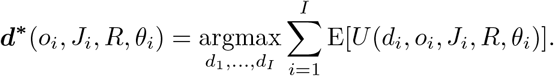

Because each term in the summation is only a function of the expected utility for one single provider *i*, this can be written as

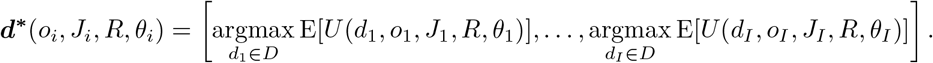

In other words, the optimal decision rule is equal to the assemblance of all individually optimal decisions, which is why we concentrate on the expected utility of a single provider hereafter.

### Consequences of classification decisions

Besides the specification of a prior distribution for *θ* and the choice of a target value *R*, the specification of a utility function is a central aspect for deriving an optimal decision rule. For the definition of utilities associated with different decisions, a time period for which decision consequences are assumed to have an effect needs to be defined. For our application to the two pathways, we assume that profiling of hospitals is carried out on a yearly basis and therefore consider the consequences of the decision to be effective only until the subsequent classification. Here and in the following, the term *utility* is used as a generic term for all positive effects of a decision compared to the effects of competing decisions, including financial consequences, transparency, good quality (or improvement) of health care, and so on.

According to the influence diagram (Fig. 1), the utility *U* may depend on the case load *J*, the parameter *θ*, the reference value *R* and the decision *D*. It is convenient to represent *U* as a 2 × 2 matrix with rows corresponding to the decision *D* and columns distinguishing whether *θ* lies below or above *R*.

| | | $\theta \leq R$ | $\theta > R$ |
| --- | --- | --- | --- |
| Classification | Non-Outlier | $\tilde{U}_{0,0}(J, \theta, R)$ | $\tilde{U}_{0,1}(J, \theta, R)$ |
| | Outlier | $\tilde{U}_{1,0}(J, \theta, R)$ | $\tilde{U}_{1,1}(J, \theta, R)$ |

Therefore, the optimal decision, which optimizes the expected utility, is given by:

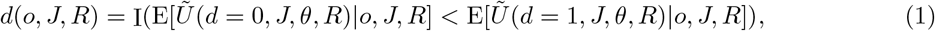

where I(·) is a 0-1 indicator function, where the value 1 is interpreted as the decision *outlier*. Following from that, it is mainly the difference *Ũ*(*d* = 1, *J, θ, R*) − *Ũ*(*d* = 0, *J, θ, R*) that is important for the classification decision. In other words, when arguing about the optimal decision rule, we may normalize the utility functions and assume that within each column of the above matrix, one of the two entries is equal to zero. Usually, if the reference value *R* is well aligned with the classification problem, the utility of a correct classification should be larger than the utility of a wrong classification.

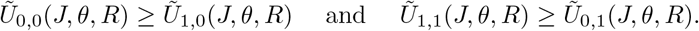

Together with the normalization, we obtain a utility matrix of the following form:

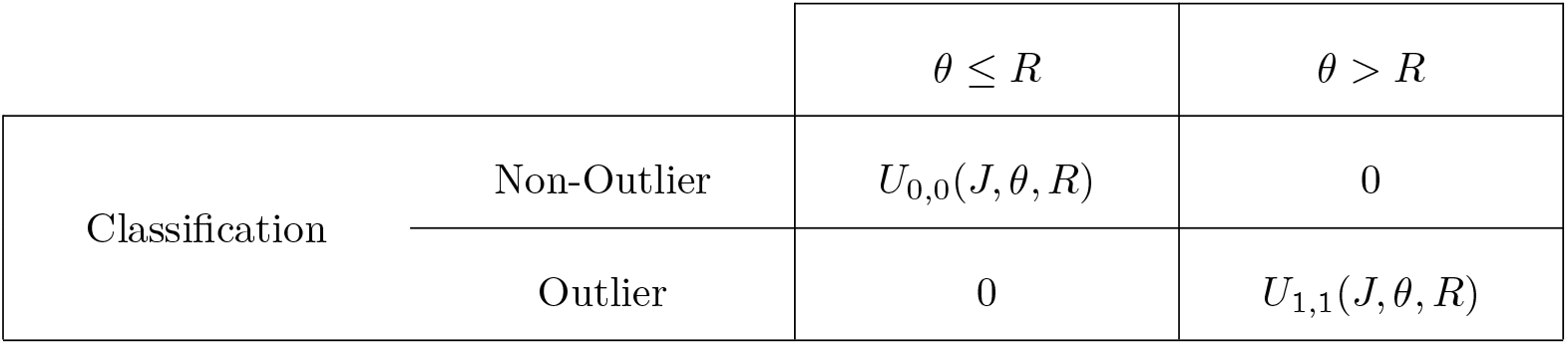

where *U*_0,0_ and *U*_1,1_ should be positive, at least when *θ* is far away from *R*.

If *U*_0,0_ and *U*_1,1_ do not depend on *θ*, then it is possible to express the optimal decision rule (1) in terms of the posterior probability P(*θ* ≤ *R*|*o, J*):

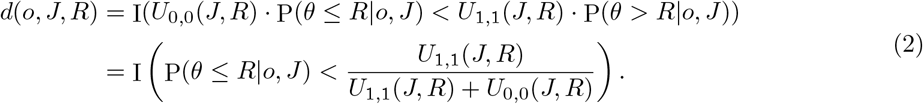

Assigning specific values to the utilities associated with classifications decisions is difficult, because the consequences are manifold and differ between the affected actors (such as patients, payers, hospitals and others). However, every decision rule either explicitly or implicitly relies on a consideration of those utilities and making those considerations explicit can lead to more transparent assessments. Below, we give examples of how the functions *U*_0,0_, *U*_1,1_ can be chosen to model simple situations. For doing so, we consider two different pathways to improve quality of care based on measurements: external reporting and change. For both, we do not aim at deriving generally valid utility functions. Instead, we illustrate how different assumptions on the functional dependencies of the utilities from *θ* and *J* affect the choice of an optimal decision rule.

### External reporting

In the pathway of external reporting, measurements of quality of care are used to reveal and compare the performance of hospitals (4). In general, external reporting and the classification of hospital performances can help to guide patient decision regarding their hospital choice (22) or support decisions for financial rewards or punishment (3,16). For simplicity, we focus in the following on external reporting with binary consequences such as accreditation of hospitals or selection for a financial reward or punishment and thus assume that the utilities are independent from the underlying quality of care of the hospital, *θ*. We allow the utilities to depend upon the number of future patients *J ′* because it is reasonable to assume that overall, decisions have a stronger effect when they affect more future patients. As described above, we are only interested in the decision consequences that have an effect until the subsequent hospital classification. Thus, *J ′* refers to the number of treated patients within the next year. For this unknown quantity, usually the current number *J* can serve as a good estimate and will be used in the following.

Thus, one possible way to define the associated utility in the pathway of external reporting is displayed in Table 1:

**Table 1:** Utility in the pathway of external reporting

| | | $\theta \leq R$ | $\theta > R$ |
| --- | --- | --- | --- |
| Classification | Non-Outlier | $\eta \cdot J$ | 0 |
| | Outlier | 0 | $\mu \cdot J$ |

with *µ, η* ≥ 0. As described above, *U*_0,0_ = *η* · *J* represents the utility of a correct non-outlier classification relative to that of an incorrect outlier classification. Thus, utilities arising independently of the classification decision (like the costs of collecting and analyzing data) are not included in *U*_0,0_

According to (2), the optimal decision rule is given by

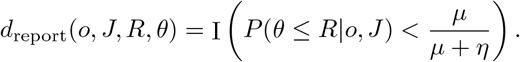

It is straightforward to show that the decision rule that we derive below is also optimal in the case in which *U*_0,0_ and *U*_1,1_ depend on *J* in the same functional way, in the sense that the ratio *U*_0,0_*/U*_1,1_ is independent of *J*. Therefore, we omit *J* as factor from Table 1 in the following.

Thus, the optimal decision rule weighs the probability of the healthcare provider being a performance outlier against the relation of expected utilities *µ/*(*µ* + *η*), in the following referred to as *α*. If, for instance, hospitals are classified as performance outliers if the probability of them being an outlier is less than *α* = 5%, this corresponds to a utility ratio of correct outlier to correct non-outlier classifications of 1 to 19 (13).

### Change in care delivery

In the pathway of change, classifications of hospitals usually intend to target resources for improvement and change processes for those providers where they have the most impact. Because the goal is to improve the quality of care of the hospitals under consideration, those who benefit most from change are future patients treated by those hospitals. In order to quantify the expected benefit, assumptions about the effects of change are necessary. These can either be made based on the evaluation of earlier actions to improve the quality of healthcare or, if no prior information is available, have to be based on a best guess. If no prior evidence is available, we suggest the following: We assume that measures of change can align the parameter of interest *θ* to the target value *R*, resulting in the improvement factor (*θ* − *R*)_+_:= max(0, *θ* − *R*). Thus, a correct outlier classification (compared to the false classification as a non-outlier) is associated with a utility depending on the potential for improvement through change, the number of patients that benefit from these improvements and some multiplication factor *δ*: *δ* · (*θ* − *R*)_+_ · *J ′*, with *δ* ≥ 0. Again, we use *J* as an approximation for *J ′*. The utility of the classification as non-outlier (compared to as outlier), on the other hand, consists of the resources that are saved given that no measures of change are implemented (denoted as *ω*). This applies regardless of whether the classification was correct or incorrect.

Following from that, one reasonable way to quantify the utilities associated with the classification decision for the pathway of change is displayed in Table 2, in which the utility of a correct outlier classification includes the resources that are necessary for the implementation of the measures of change (*ρ*):

**Table 2:**
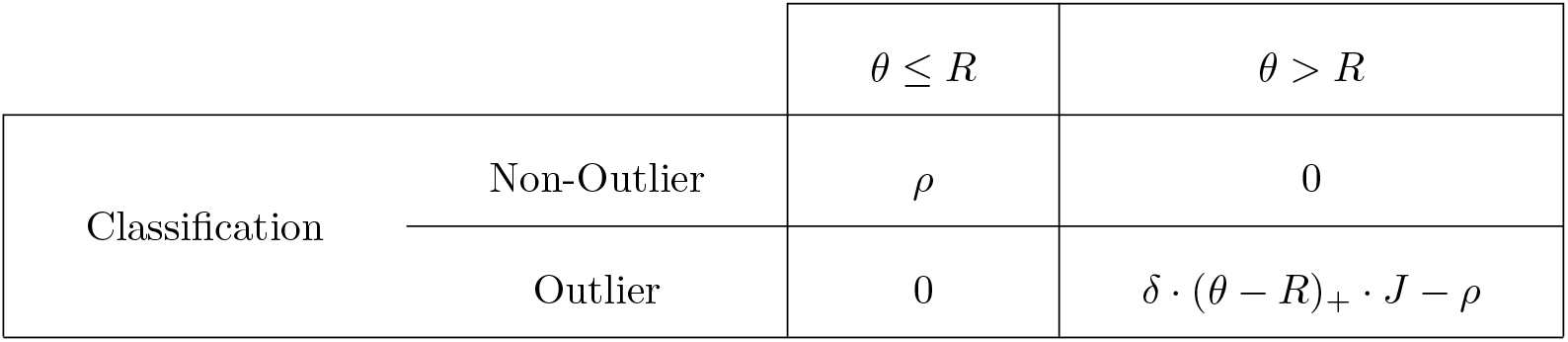
Utility in the pathway of change

| | | $\theta \leq R$ | $\theta > R$ |
| --- | --- | --- | --- |
| Classification | Non-Outlier | $\rho$ | 0 |
| | Outlier | 0 | $\delta \cdot (\theta - R)_+ \cdot J - \rho$ |

with *ρ, δ* ≥ 0. Based on this, the optimal decision rule can be derived as:

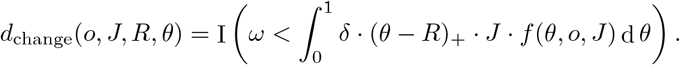

Again, not the absolute utilities but only the relation of *ω* and *δ*, in the following referred to as *ω* = *ρ/δ*, has to be defined in order to derive a utility-efficient decision rule. Here, *ω* can be interpreted as the relation of utilities between those of saving resources by not implementing measures of change for one hospital and those of its future patients benefiting from measures of change. In contrast to the pathway of external reporting, the utilities of change are assumed to be directly dependent upon the outcome probability *θ*. In this case, it is not possible to give a simple expression for *d*_change_ as a function of posterior probabilities.

### Comparison of different decision rules

In order to derive optimal decision rules *d*_report_ and *d*_change_, the expected utilities associated with a classification of a hospital as performance outlier have to be set into relation to those associated with a classification as non-outlier. For the paradigmatic utility functions for the two pathways described above, this is controlled by the two parameters *α* and *ω*, respectively. Given those and a fixed number of treated patients *J*, critical values *o*_min_ for the observed number of outcomes, *o*, can be calculated which lead to the classification as performance outlier:

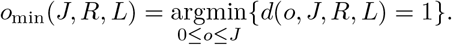

Hospitals with observed *o* ≥ *o*_min_ are classified as performance outliers, where the concrete value of *o*_min_ is pathway-specific.

To illustrate the effects of the hospital volume (*J*) and the choice of a decision rule (*d*_report_ and *d*_change_), the critical values *o*_min_ are displayed in a funnel plot (5). As an example, we use a exemplary quality indicator with a target value (up to) *R* = 15%. Because we assume no prior knowledge about the parameter, we use the non-informative Jeffrey’s prior as prior Beta(1*/*2, 1*/*2) distribution for *θ*.

In addition to the decision rules *d*_report_ and *d*_change_, we analyze the decisions resulting from a naive decision rule:

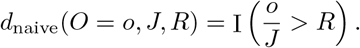

This naive rule has been traditionally used for classification decisions in Germany (23), which is why we compare its results to that of the decision rules described above.

We compare the three decision rules with respect to their critical values *o*_min_ as well as their sensitivity and specificity in three scenarios: with underlying quality of care within (*in control*: *θ* = 10%), outside (*out of control*: *θ* = 20%), and at the border of the reference area (*borderline*: *θ* = *R* = 15%). Sensitivity is here defined as the probability of identifying outlying hospitals as performance outliers (and applies thus only to the out-of-control scenario), and specificity as probability of identifying non-outliers as non-outliers (and applies to the in-control and borderline scenario). For each scenario, sensitivity and 1 − specificity are calculated for a varying number of patients treated by the hospital. Again, Jeffrey’s prior is used.

We apply the above proposed decision rules to five quality indicators covering aspects of stationary hip replacement surgeries in Germany in the year 2017. For all hospitals, providing relevant services for these indicators, we calculate for each indicator the proportion of hospitals which are classified as performance outliers given various values for *ω* and *α* and using Jeffrey’s prior for *θ*. Because in Germany, *d*_naive_ is currently used as decision rule and all classifications of performance outliers are subsequently validated in a qualitative step, we can compare the positive-predictive-value (PPV) of the proposed decision rules for various values of *ω* and *α* with respect to this qualitative assessment. The validation consists of a structured dialogue between the hospital and the responsible quality assurance agency and aims at determining whether the classification as outlier is really due to deficits in the quality of care (1,2). PPV is calculated as the proportion of hospitals whose classification as performance outlier is confirmed by the qualitative validation.

## Results

Figure 2 shows the critical values *o*_min_ depending on *J* for an indicator with *R* = 15% and for specific values of *ω* and *α*:

**Figure 2:**
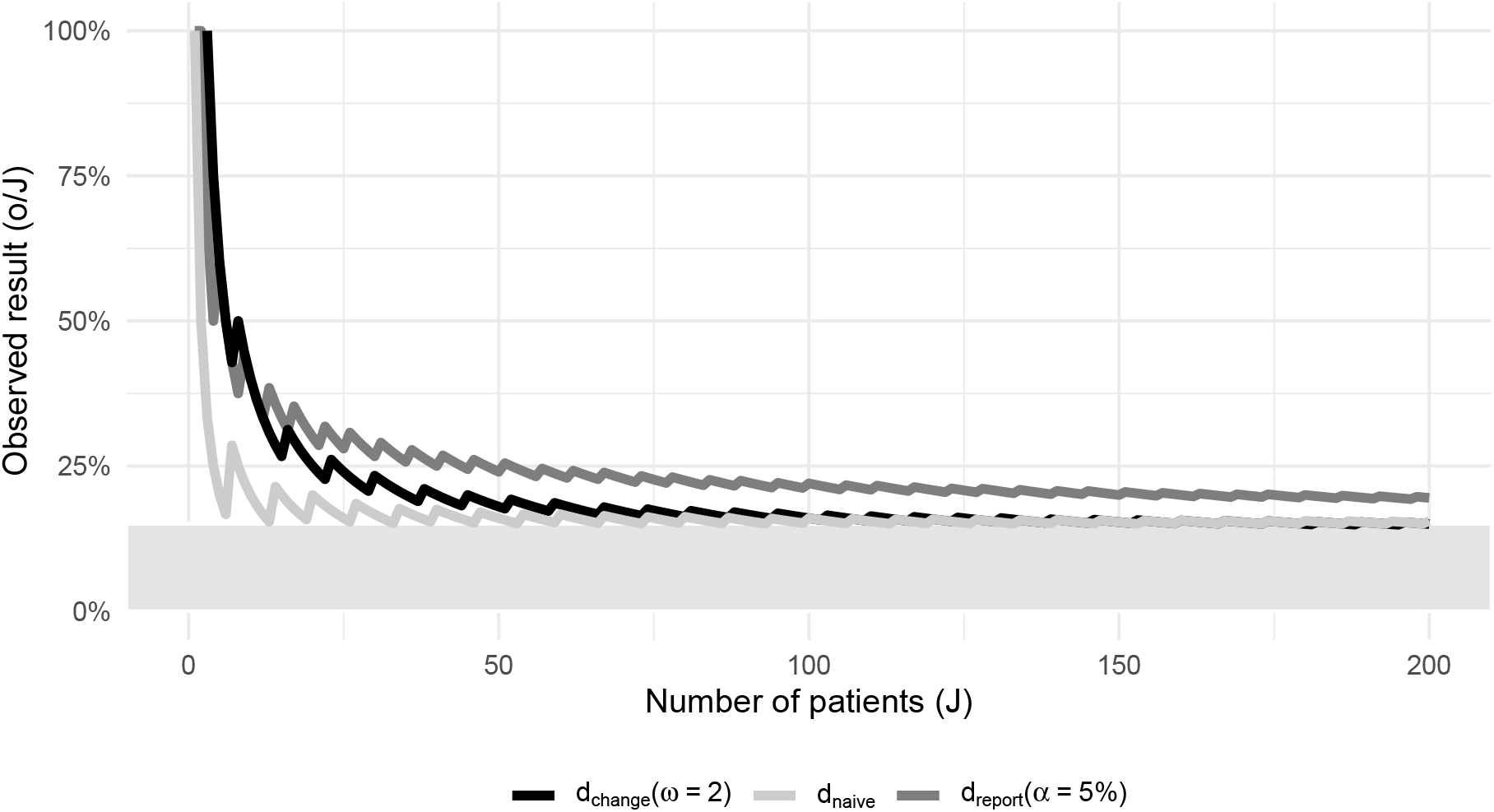
Funnelplot for a binary indicator with *R* = 15%

With increasing number of patients *J*, the critical value of all three decision rules approaches the target area (grey area in Figure 2). For low-volume providers, *d*_change_ leads to more liberal decisions compared to *d*_report_, with *d*_naive_ yielding the lowest critical values. On the other hand, *d*_change_ converges more quickly to *R* than *d*_report_ for higher values of *J*. This means that for the specific values *α* = 5% and *ω* = 2, low-volume hospitals are more often classified as performance outliers based on *d*_report_ compared to *d*_change_, while the opposite is true for high-volume hospitals. The critical value of *d*_naive_, in turn, is the most conservative over almost all *J*.

This is also reflected in the sensitivity and specificity of all three decision rules displayed in Figure 3:

**Figure 3:**
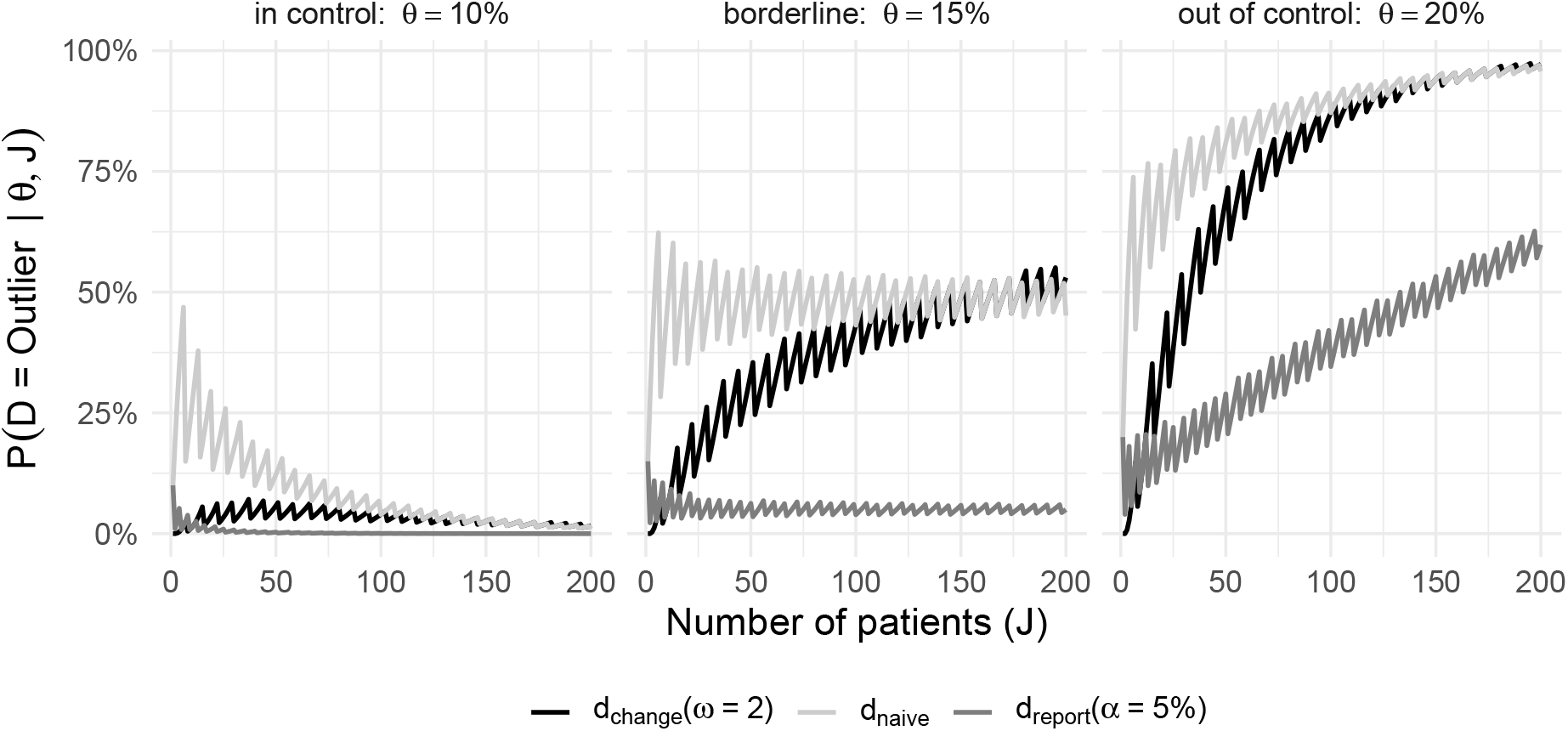
Sensitivity and specificity as a function of the number of cases *J*

For the in-control-scenario, the probability of a false-positive classification are low for both *d*_report_ and *d*_change_, indicating a high specificity. For *d*_naive_, however, the specificity is comparatively low for small values of *J*. For the borderline-scenario, the decision rule *d*_report_ for the pathway of external reporting has, on average, a probability of 5% for false-positive classifications, corresponding to a specificity of 95%. The decision rule for the pathway of change, *d*_change_, has a higher specificity for low-volume providers. With increasing number of patients, however, the probability of false-positive classifications increases, due to the fact that the expected utility of false-negative classifications increase with *J*. This, on the other hand, means that *d*_change_ has a high sensitivity for high-volume providers, which can be seen in the out-of-control-scenario. There, the sensitivity of *d*_report_ also increases with higher numbers of patients, but not as quickly as that of *d*_change_. For low-volume providers, however, the probability of false-negative classifications is again higher for *d*_change_. For *d*_naive_, the specificity is low if the true parameters lies within the reference area, but the sensitivity is high even for small-volume providers compared to both other decision rules if the true parameter lies outside of the reference area.

The above figures only show the consequences of single choices *α* = 5% and *ω* = 2 for the parameters, but the results are qualitatively similar for other values. Consequences of alternative choices for the parameters can be investigated via two Shiny apps (24).^2^ Also for different choices of *α* and *ω*, the general conclusion holds that for the pathway of change, outlier classifications are more frequent for high-compared to low-volume providers compared to the pathway of external reporting, with similar consequences for both sensitivity and specificity as presented above.

### Application to hospital profiling in Germany

In total, 1,277 hospitals were profiled based on data from more than 180,000 treated patients in 2017 for the five considered quality indicators for hip replacement surgeries (25). An overview over all indicators is given in Table 3:

**Table 3:** Quality indicators for hip replacement surgeries

| QI | Quality indicator | Reference area |
| --- | --- | --- |
| 1 | Patient has no indication for elective hip replacement surgery | $\leq 10\%$ |
| 2 | Patient has no indication for hip revision surgery | $\leq 14\%$ |
| 3 | Length of stay prior to surgery for hip fracture is $> 48\text{h}$ | $\leq 15\%$ |
| 4 | No measures for fall prevention taken | $\leq 20\%$ |
| 5 | Patient is not mobile on discharge | $\leq 5\%$ |

Over all hospitals and indicators, 851 out of 5,270 hospital results were reported as being classified as performance outliers based on the naive decision rule *d*_naive_. In the qualitative validiation of those outlier classifications, only 17% were subsequently confirmed as actual outliers.

For all hospitals and quality indicators, the number of hospitals that would have been classified as outliers using *d*_report_ and *d*_change_ is displayed for various values of *ω* and *α* in Figure 4:

**Figure 4:**
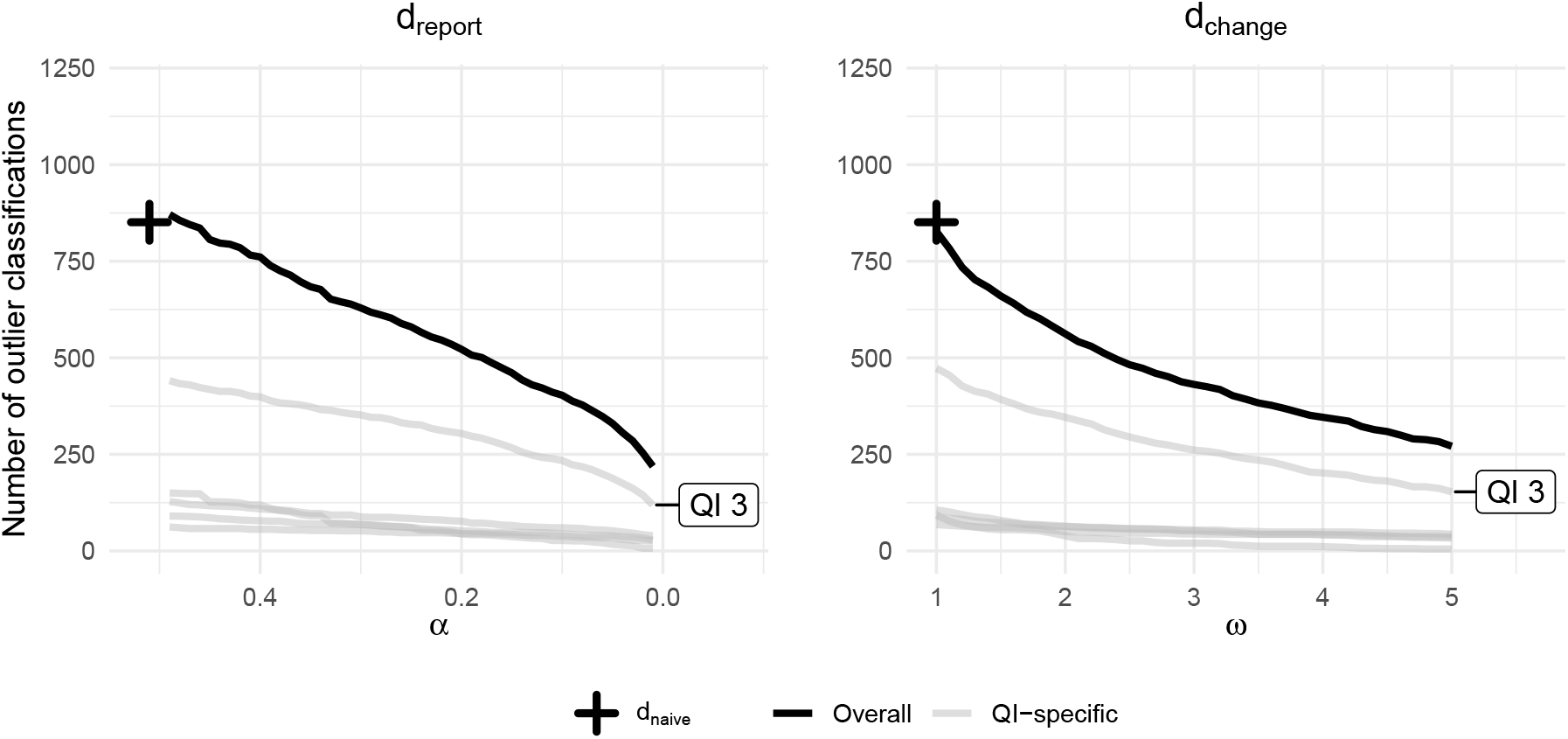
Number of outlier classifications as functions of the parameters *α* and *ω*

For all considered values, the naive decision rule is the most conservative, yielding the highest number of outlier classifications (851). For the two other decision rules, the number of outlier classifications decreases with decreasing *α* and increasing *ω*, respectively. This is especially true for QI 3 (length of stay prior to surgery), for which the number of outlier classifications is generally the highest among all indicators. For all other indicators, the number of outlier classifications is lower but decreases similarly with decreasing *α* and increasing *ω*, respectively.

Figure 5 displays the PPV, that is the proportion of outlier classifications that were confirmed as outliers in the qualitative validation step:

**Figure 5:**
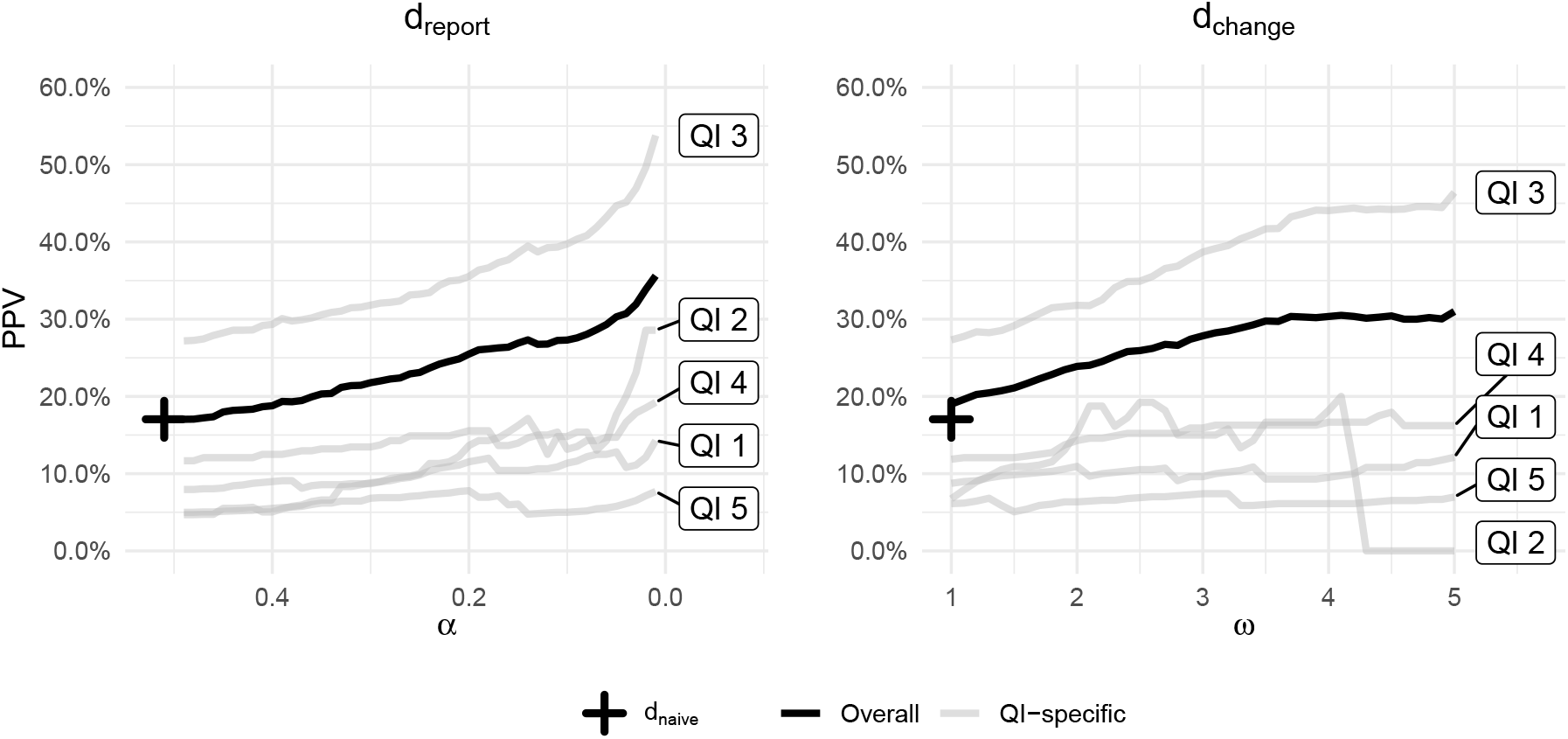
PPV as functions of the parameters *α* and *ω*

For both decision rules, the overall PPV is low for large values of *α* and low values of *ω* but increases to up above 30% over all indicators and for both *d*_report_ and *d*_change_. Indicator 3 (length of stay prior to surgery) is the indicator for which the PPV increases the most, from about 25% for *ω* = 1 and *α* = .5 and up to and above 50% for *ω* = 5 and *α* = .01. For all other indicators, there is at most a weak relationship between the PPV and the values of *α* and *ω*.

Overall, even with conservative decision rules that assume much lower utility for false-positive compared to false-negative classifications, the PPV is relatively low across all hospitals and quality indicators. Only the PPV of a single indicators shows a strong relationship between the PPV and increasing or decreasing values of *ω* and *α*, respectively.

## Discussion

The classification of hospitals on the basis of measurements of process or outcome indicators is a decision under uncertainty, because the observed data only allows probabilistic statements about the true parameter of interest. Bayesian decision theory provides a framework to derive decision rules that are optimal with respect to the expected utility associated with each decision. Because the utilities are a crucial part of the decision rule, they should be reflected upon and incorporated into the decision rule wherever possible in order to make decisions transparent and justifiable. Funnel plots prove to be an intuitive way to communicate the resulting decision boundaries. In practice, however, arriving at a single utility function can be difficult, especially because consequences for different parties (such as patients, hospitals, sponsors, investigators, policy makers) have to be considered and weighted against each other. In these cases, it can be useful to define a set of fixed decision rules (based on values such as *α* ∈ {0.01, 0.05, 0.1, 0.2, 0.4} and some integer values of *ω*, respectively) and, by doing so, to contrast different underlying utilities in order to arrive at a rule that suits best to the decision problem at hand among a set of options.

For the simultaneous classification of multiple hospitals, we assumed an additive utility function. In practice, the assumption of simple additivity may not always be a appropriate. For instance, the quality and effects of measures of change can be dependent upon the total number of outlier classifications due to limited resources. In this case, the utility of an outlier classifications directly depends upon the overall number of outlier classifications, which should be incorporated into the utility function. Furthermore, we ignored that the decision rule itself can have an effect on the quality of care provided by hospitals. If, for instance, small-volume hospitals below a certain number of treated cases irrespectively of their performance are never classified as outliers, the absence of quality assurance at these hospitals could lead to a decline in the quality of care. In this case, it can be reasonable to modify the decision rule in order to avoid undesired effects or use additional instruments.

The application of the presented framework to the German setting of hospital profiling in the area of hip replacement surgeries illustrated that the chosen utility function has a high impact on the number of outlier classifications: the lower the utility of correct non-outlier in relation to correct outlier classifications are assumed to be, the more providers are classified as performance outliers. One consequence of a low utility for correct non-outlier in relation to correct outlier classifications (resulting in large values for *α* and small values for *ω*, respectively) is a lower PPV of the classification decision. However, the relationship between PPV and values of *α* and *ω* was pronounced only for one of the five quality indicator and weak or non-existent for all others. This is surprising, because one could assume that more liberal threshold values should lead to a higher PPV among those still classified as outliers. One reason for the weak relationship between PPV and values of *α* and *ω* could be that the validation of results classified as outliers is a relatively informal process with plenty of heterogeneity and, hence, cannot serve as a gold standard for the classification of true outliers (23). Also, it is possible that the indicators’ inclusion criteria are not accurate enough to exclude cases from the indicators which are explained by special circumstances. Our suggested explicit addressing of uncertainty as part of the decision underlines that the weak relationship between the PPV and *α* and *ω* could be reason to review the validity of the indicators or the qualitative validation process.

Profiling of hospitals needs to be based upon state-of-the-art statistical methodology. Our work offers a flexible framework, which can can easily be extended to other types of quality indicators, including risk-adjusted indicators, the aggregation of several indicators or a sequential decision making context.

## Conclusion

The classification of hospitals on the basis of measurements of process or outcome indicators is a decision under uncertainty and can be analyzed using a decision-theoretic framework. This allows the contrasting of different decision rules regarding their underlying assumptions and consequences. Funnelplots and the analysis of sensitivity and specificity of decision rules are useful ways to examine their characteristics and consequences, also with regard to undesired side effects. This way, the effectiveness and transparency of decision making in the context of hospital profiling can be improved.

## Data Availability

The data are collected and analyzed as part of the mandatory routine external quality assurance program in Germany based on paragraph 136ff SGB V. The data are part of the public reporting occuring under the rules of paragraph 137 Abs. 3 Satz 1 Nr. 4 SGB V (Qualitaetsbericht der Krankenhaeuser; Qb-R) for the reporting year 2017. The quality reports of the hospitals are only used in part. A complete, unchanged representation of the quality reports of the hospitals is available at https://www.g-ba.de, where data usage can also be requested.

## Declarations

### Ethics approval and consent to participate

The data are collected and analyzed as part of the mandatory routine external quality assurance program in Germany based on §136ff SGB V. Thus, no ethical approval of the study was necessary.

### Consent for publication

Not applicable

### Availability of data and materials

The data are collected and analyzed as part of the mandatory routine external quality assurance program in Germany based on §136ff SGB V. The data are part of the public reporting occuring under the rules of § 137 Abs. 3 Satz 1 Nr. 4 SGB V “Qualitätsbericht der Krankenhäuser” (Qb-R) for the reporting year 2017.^3^ The quality reports of the hospitals are only used in part. A complete, unchanged representation of the quality reports of the hospitals is available at https://www.g-ba.de, where data usage can also be requested.

### Competing interests

The authors declare that they have no competing interests

### Funding

Not applicable

### Contributions

JH: made substantial contributions to the conception, analysis, interpretation of results, and has drafted the work. JR and JC: made substantial contributions to the interpretation of results and have substantively revised the work. MK: made substantial contributions to the analysis and has substantively revised the work. MH: made substantial contributions to the conception and design of the study, the interpretation of results and has substantively revised the work. All authors read and approved the final manuscript.

## Acknowledgements

Not applicable

## Footnotes

1 External reporting is sometimes also referred to as *accountability* or *selection* and change as *improvement* (3,4)

2 https://iqtig.shinyapps.io/funnel_plot/ and https://iqtig.shinyapps.io/sensitivity_specificity

3 https://www.g-ba.de/richtlinien/39/

## Notes

### Competing Interest Statement

The authors have declared no competing interest.

### Funding Statement

No external funding was received.

### Author Declarations

The data are collected and analyzed as part of the mandatory routine external quality assurance program in Germany based on paragraph 136ff SGB V. Thus, no ethical approval of the study was necessary.

